# Socioeconomic inequity in access to care among older people in Japan

**DOI:** 10.1101/2023.06.12.23291290

**Authors:** Shohei Okamoto, Atsuhiro Yamada, Erika Kobayashi, Jersey Liang

## Abstract

**Background:** Equity in access to long-term care (LTC) enables older people to maintain their well-being after they undergo a decline in their intrinsic capacity.

**Methods:** We used data from Wave 6 (2002) through Wave 10 (2021) of the National Survey of the Japanese Elderly to assess gradients by income and education in access to medical care and LTC among Japanese individuals aged 60 years and above. Specifically, we assessed self-reported unmet needs for medical care and LTC, and public LTC use, and estimated the concentration indices (CI) to measure the degree of inequality and inequity. We standardised public LTC use by need and non-need variables. We analysed data derived from up to 1,775 person-wave observations from 1,370 individuals.

**Findings:** The pooled incidence across waves of forgone medical care, self-reported unmet support for activities of daily living (ADL) or instrumental ADL (IADL), and those not certified for LTC services even with ADL or IADL limitations were 4.6%, 15.5%, and 62.5%, respectively. Public LTC use demonstrated pro-higher education and pro-rich distribution, whereas the gaps decreased for need-predicted use. Based on the CI estimates, no explicit inequality was found for forgone medical care. However, we observed inequity in standardised LTC use across education, indicating pro-higher education inequality, particularly among women and those aged ≥80 years.

**Conclusion:** Improving the understanding of available resources and strengthening the functions of health centres and communities are required to detect the needs of citizens and facilitate their access to necessary care.

## Background

Universal health coverage (UHC) means everyone, everywhere can access nationally determined sets of needed essential health services, including promotion, prevention, treatment, rehabilitation, and palliative care without incurring financial hardship (1). Older people may require more care to maintain their functional ability (i.e. the interaction of intrinsic capacity with environmental characteristics), human dignity, and well-being because their intrinsic capacity (i.e. a combination of their physical and mental capacities) declines (2). Ageing population is a global trend (3); therefore, ensuring equitable access to necessary care, including long-term care (LTC) for older people, is essential to achieve progress toward UHC.

LTC includes numerous services required by people dependent on assistance with the basic activities of daily living (ADL) (4). Among the OECD countries, the total LTC expenditure per country’s gross domestic product ranges from 0.1% to 4.1%, in addition to differences in care providers (e.g. nursing homes, hospitals, home care, households, and social providers) (5). To date, only six countries have adopted national social LTC insurance systems, namely the Netherlands, Israel, Germany, Luxembourg, South Korea, and Japan (6). Japan is the first Asian country, which has adopted a public universal LTC insurance in 2000, principally covering all citizens aged ≥40 years (7). To be eligible for public LTC benefits, people requiring care undergo an objective test to evaluate their level of ADL dependence and an assessment by their attending physician and are classified into independence, support-need-levels 1 to 2, and LTC need-levels 1 to 5. The types and amounts of available benefits are determined based on this classification. The users bear charges for LTC services at a 10% co-payment rate (or 20% or 30% depending on one’s income level). In addition, several financial protection policies are available to mitigate excessive out-of-pocket payments and the burden of insurance premiums for low-income individuals. Further details on LTC insurance in Japan have been described previously (7).

### Socioeconomic inequity in access to care and unmet LTC needs

Despite evidence of socioeconomic inequity in access to healthcare (8–10), there is limited evidence of such inequity in LTC. Based on the Grossman model (11), socioeconomic inequity in access to healthcare can arise from various channels, such as disparities in health literacy, income, time preference, and available time for producing health. However, inequitable access to LTC can be driven by different channels from healthcare, including due to substitution between formal and informal care. Those unable to access the necessary care experience unmet needs, which could lead to negative health outcomes (12, 13); or ensuring access to LTC can have a protective effect for increased medical care expenditures, such as one for emergency care use (14). Women, lower education, and worse economic status predict unmet needs for personal assistance in ADL among older adults (15, 16). Inequity in LTC access exists in South Korea and the US, suggesting the importance of extending insurance coverage and subsidising low-income individuals (17–19). Formal care among people without dementia is pro-rich, and poor people are more likely to experience unmet care needs in England (20). Furthermore, findings from Spain and the Survey of Health, Aging, and Retirement in Europe suggest that formal services are concentrated among richer people, whereas poorer people tend to use intensive informal care (21, 22). Studies in the Netherlands also find a pro-poor gradient in home care use (23, 24). However, a Japanese study found that formal LTC use was generally equitable across income groups (25).

LTC services may still exhibit economic inequity despite the availability of universal insurance or benefits, as suggested by studies in South Korea and European countries (17, 18, 22). Nevertheless, income inequity in LTC service use among those eligible for LTC benefits has not been observed in Japan (25), potentially owing to its low co-payment rate and financial protection policies. However, as mentioned before, socioeconomic inequity in care access can arise from financial and non-financial causes. Under the social insurance scheme, users need to contract with providers to purchase services at a (quasi) market, which would require a certain level of knowledge about LTC services and their own preferences in service use. Furthermore, in Japan, individuals are required to submit an application to be eligible for public LTC services, which may serve as a hindrance to accessing care for non-financial reasons. Therefore, we aimed to expand the literature in assessing socioeconomic inequity in LTC access using both financial and non-financial indicators under the public universal LTC insurance system.

### Methods

#### Data

Data were obtained from the National Survey of the Japanese Elderly (26), comprising Japanese adults aged ≥60 years. The survey was first conducted in 1987 and was followed up with participants every 3 to 6 years, adding new samples to complement sample size declines caused by deaths and a loss to follow-up. The sample was extracted from the Basic Resident Registration System using a stratified two-stage random sampling method based on a combination of regional blocks and population. From waves 1 to 10, 7,892 individuals responded in one or more waves. To evaluate access to LTC services, we analysed data collected from wave 6 (2002) to wave 10 (2021), after public LTC insurance introduction in 2000.

We restricted our analysis to participants who responded to interviews, excluding mail, proxy, and non-responders’ surveys because LTC-need-related information and self-reported unmet needs were only available for these people. Thus, we obtained 10,743 person-wave observations from 5,471 unique, non-institutionalised individuals from Wave 6 to Wave 10. We then focused on those with LTC needs, leading to 1,975 person-wave observations by 1,512 individuals. Finally, we excluded those with missing information, resulting in the final sample size of at most 1,775 person-wave observations by 1,370 individuals.

#### Access to medical care: Forgone care

To assess the inequality in medical care access, we used self-reported forgone medical care (10, 27). In the survey, for the question, ‘During the past 3 months, how often did you reduce the dose of medication or did not see a physician even though it was necessary?’, each respondent selected one of the following options: 1. most times; 2. sometimes; 3. hardly/none; and 4. they did not need to consult a physician or consume medicine. Respondents who selected ‘most times’ or ‘sometimes’ were regarded as forgoing needed care. We excluded those who did not need to consult a physician or consume medicine. To assess inequality in medical care access, we analysed the data obtained in waves 8, 9, and 10, because the questionnaire was administered only in these waves.

#### Access to LTC

##### (1) Self-reported unmet needs

Respondents with LTC needs were questioned if someone else (e.g. family members and LTC workers) had helped them with their activities within the past 3 months. They rated the frequency of receiving support as follows: 1. almost always; 2. sometimes; 3. occasionally; 4. never; and 5. did not need support. Respondents who selected ‘sometimes’, ‘occasionally’, or ‘never’ were categorised as having experienced an unmet need for LTC. This operational definition is reasonable because people who cannot obtain necessary care whenever they need them experience unmet need.

To define the LTC needs operationally, we used questionnaires on difficulties in performing basic and instrumental activities of daily living (ADL and IADL). Similar to the Katz index (28), ADL was measured using the following six items: bathing, dressing, feeding, transferring, outing, and toileting. IADL was measured using the following four items: shopping for personal items, using a telephone, riding the bus or subway alone, and performing light tasks around the house (29). Each item was rated on a five-point Likert scale, ranging from 0 (never difficult) to 4 (unable to do at all). Respondents experiencing difficulties with at least one of the items (i.e. 2. moderately difficult to 5. unable to do it at all) were defined as having LTC needs.

##### (2) Utilisation-based measurement

Considering health disparities across socioeconomic statuses (30, 31), the care need differs across groups, leading to dissimilar levels of care utilisation. Thus, higher utilisation among the poor with worse health status does not imply pro-poor inequity in care access based on the vertical equity principle. Therefore, standardised utilisation must be used to assess inequity in access to care (32).

We standardised LTC use by the LTC need explained by demographics, health status, and morbidity status (32), formalised as a non-linear model with repeated measures (i.e. multilevel mixed-effects logistic regression) as follows:

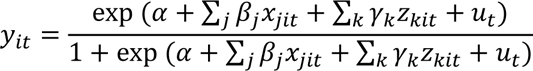

where *y*_*it*_ denotes a binary variable corresponding to 1 if respondent *i* in year *t* is considered eligible for public LTC services at any level, and 0 otherwise. The *j*th need variables (*x*) to predict LTC use and *k*th control variables (*z*) to avoid omitted-variables bias, with parameters *β* and *γ*, are included. α is a constant, and *u* is the random effects. Accepting that the variance of need-standardised use may depend on the formulation of the *z* variables in the standardisation procedure, need-standardised LTC use (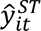) is defined as:

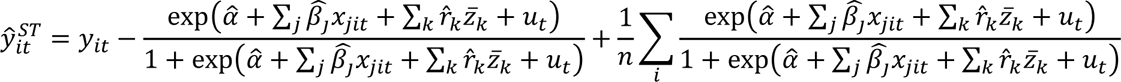

where *n* denotes the sample size, and the *z* variables are set to their means (*z̅*). The need variables directly influencing LTC needs are proxied by the demographic and morbidity characteristics (32), including the age, sex, self-rated health, the number of chronic conditions (ranging from 0 to 6), the degree of difficulty in performing each ADL (0–24) and IADL (0–16), and cognitive functioning (0–9). The non-need control variables include the marital status, the number of co-resident members, residential area, the population size of the residential area, and year dummies. These non-need variables may affect the health status through social support and relationships (33); however, they are expected to affect formal and informal care availability, generating potential heterogeneity across group behaviour in the application and utilisation of formal LTC services. The need and non-need variables are defined in the Appendix Table A-1.

#### Empirical strategy

##### Concentration index

To measure inequality and horizontal inequity in the access to medical and LTC services across socioeconomics groups, we estimated the concentration indices (CI) frequently used to evaluate health disparities (32). CI in each year is defined as twice the area between the concentration curve and the 45° line as follows (32):

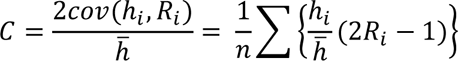

where ℎ_*i*_ denotes the health variable of interest to measure inequity, and *ℎ̅* is its mean. *R*_*i*_ is the rank variable in which the health gradients are measured. For binary outcomes, the concentration index (W) was rescaled to consider its dependence on the variable means and to satisfy the mirror condition, i.e. the absolute value of a measured inequality is symmetric when computed over either attainments or shortfalls as follows (34):

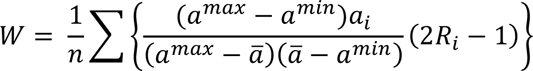

The CI is positive when access is more frequent among higher socioeconomic groups and negative if otherwise. We used the years of education and income as the rank variables to measure inequality and inequity. For income, we used the couple’s gross annual income equalised by the marital status (i.e. divided by the square root of 2 if married) and residualised by employment status (See Appendix B). To calculate the descriptive differences in LTC utilisation across education and income groups, but not the CI, both indicators were divided into five categories. The category for education included lowest (<6), lower-middle (6–9), middle (9–12), higher-middle (12–15), and higher (16+). Despite different pre- and post-World War II educational systems, this categorisation roughly corresponded to the distinctions between elementary, secondary, high school, and university or higher. Income was divided into 20^th^ percentile groups.

Demographic factors, such as sex and age, can influence help-seeking behaviours (35); thus, we evaluated inequities according to the sex and age. Considering that the average age of the sample was approximately 81, we categorised the participants into those aged <80 years and ≥80 years. This categorisation is reasonable because people can become drastically dependent on ADL around the age of 80 years (36).

To partially adjust for potential selection bias, we adopted two types of weights, namely, cross-sectional and longitudinal. Cross-sectional weights were estimated by logistic regression as probabilities of responding to baseline surveys predicted by the age, sex, the geographic area of residence, and the municipal/population category of a residential area. Longitudinal weights were estimated as response probabilities in each wave, estimated by the age, sex, employment status, marital status, education, self-rated health, the geographic area and population category of a residential area at baseline or the closest. These approaches were similar to multiple imputations based on random missing data (37, 38).

## Results

### Descriptive statistics

Table 1 summarises the incidence of forgone medical care and unmet LTC needs in each wave. The average incidence for medical care was 4.6% across the waves as follows: 4.5%, 6.0%, and 3.8% in 2012, 2017, and 2021, respectively. The average incidence for ADL or IADL support was 41.6%, ranging from 37.9% to 44.6% from 2002 to 2021. Approximately, 62.0% of the participants with ADL or IADL limitations were not certified for LTC services (55.3–71.8%).

**Table 1.**
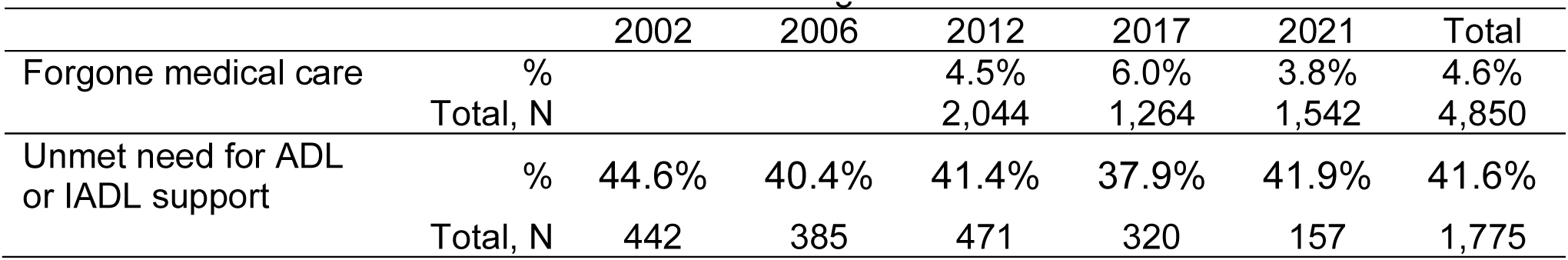
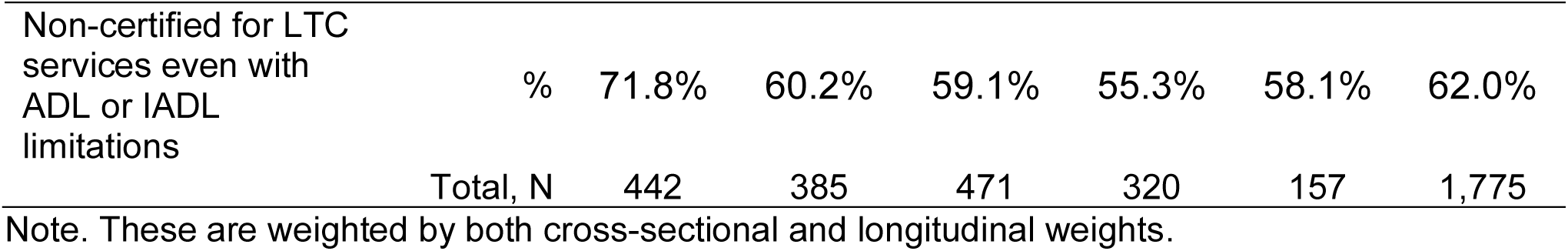
Incidence of unmet need for medical and long-term care.

Table 2 defines the pooled descriptive statistics between Wave 6 (2002) and Wave 10 (2021) for those with LTC needs. Of all participants, 38.0% were certified for public LTC use. The average age was approximately 80.8 years, and the majority of participants were women (72.6%).

**Table 2.** Pooled descriptive statistics, Wave 6 (2002) to Wave 10 (2021)

| Stats | % or mean | Standard<br>Deviation |  | % |
| --- | --- | --- | --- | --- |
| Certified for long-term care use | 38.0% |  | Residential area<br>(Geographic region<br>of Japan) |  |
| Standardised long-term care<br>use | 0.38 | 0.44 | Hokkaido | 3.6% |
| Years of education | 9.28 | 2.58 | Tohoku | 10.1% |
| Couple's income (10,000JPY) | 175.21 | 152.43 | Kanto | 18.8% |
| Age | 80.80 | 6.28 | Hokuriku | 7.1% |
| Women | 72.6% |  | Tozan | 6.9% |
| Employment status: Working | 5.1% |  | Tokai | 8.0% |
| SRH: Very good | 11.5% |  | Kinki | 13.2% |
| SRH: Good | 39.8% |  | Chugoku | 9.7% |
| SRH: Fair | 35.6% |  | Shikoku | 4.3% |
| SRH: Bad | 11.1% |  | Kyushu | 18.3% |
| SRH: Very bad | 2.0% |  | Municipal/population<br>category:<br>Government-designated | 13.0% |
| N of chronic conditions | 2.02 | 0.99 | Population size:<br>200K+ | 20.0% |
| ADL score (0–24) | 2.77 | 3.87 | Population size:<br>100K–200K | 16.4% |
| IADL score (0–16) | 4.79 | 4.22 | Population size:<br><100K | 28.1% |
| Memory test (0–9) | 1.89 | 1.71 | Population size:<br>Town and villages | 22.5% |
| N of household members | 1.94 | 1.69 |  |  |
| Marital status: Single | 55.1% |  |  |  |

Note: SRH stands for self-rated health; ADL stands for activities of daily living; IADL stands for instrumental activities of daily living; government-designated cities have populations greater than 500,000 and have been designated by the Cabinet of Japan; these descriptive statistics are calculated for 1,775 respondents with long-term care needs (i.e. ADL or IADL limitations), weighted by both cross-sectional and longitudinal weights; for couples' income, the sample size is 1,375; individuals whose marital status is single include those divorced and widowed.

Appendix Table A-2 presents the regression results to predict LTC use by need and non-need variables. Both needs (e.g. chronic conditions and ADL and IADL limitations) and non-needs (e.g. the number of co-resident members) were associated with the certification for public LTC use.

Table 3 summarises the mean probabilities of non-standardised, need-predicted, and need-standardised LTC use by education and income, pooling the data between waves 6 and 10. Remarkably, the actual distribution of public LTC was pro-higher-education and pro-rich, whereas the gaps decreased for need-predicted use. Thus, the participants with lower education and income used public LTC services less than expected by 3% to 7% points, whereas their counterparts with middle and higher levels used services more than expected by 3% to 26% points. Despite standardisation, higher education and income groups used more services than the lowest groups.

**Table 3.** Non-standardised and standardised long-term care use by education and income: Mean probabilities, Pooled between Wave 6 (2002) and Wave 10 (2021)

|  |  | Actual | Need-predicted | Difference | Standardised |
| --- | --- | --- | --- | --- | --- |
| Education | Lowest: <6 | 0.30 | 0.37 | -0.07 | 0.31 |
|  | Lower middle: 6–9 | 0.37 | 0.39 | -0.03 | 0.35 |
|  | Middle: 9–12 | 0.46 | 0.38 | 0.08 | 0.45 |
|  | Higher middle: 12–15 | 0.42 | 0.39 | 0.03 | 0.40 |
|  | Higher: 16+ | 0.70 | 0.44 | 0.26 | 0.64 |
| Income<br>(Quintile) | Poorest 20% | 0.34 | 0.37 | -0.03 | 0.34 |
|  | 2nd poorest 20% | 0.35 | 0.38 | -0.02 | 0.35 |
|  | Middle | 0.42 | 0.38 | 0.04 | 0.42 |
|  | 2nd richest 20% | 0.49 | 0.38 | 0.10 | 0.48 |
|  | Richest 20% | 0.42 | 0.37 | 0.05 | 0.42 |

### Socioeconomic inequality and inequity in access to medical care and LTC

Table 4 summarises CIs for medical care and LTC use by education and income in each wave. Figure 1 presents visualised horizontal inequities in LTC use by education and income. We observed no explicit inequality for forgone medical care. However, inequity was observed in LTC use across education in 2006 and 2012, indicating pro-higher-education inequality: CIs with standard errors were 0.12 (0.03) and 0.09 (0.03). Meanwhile, the inequity was less obvious for income, showing that concentration curves crossed 45-degree lines in most waves and pro-rich inequality was evident only in 2012. For non-need-standardised self-reported unmet needs for LTC support, both education and income inequality were evident in 2002.

**Figure 1.**
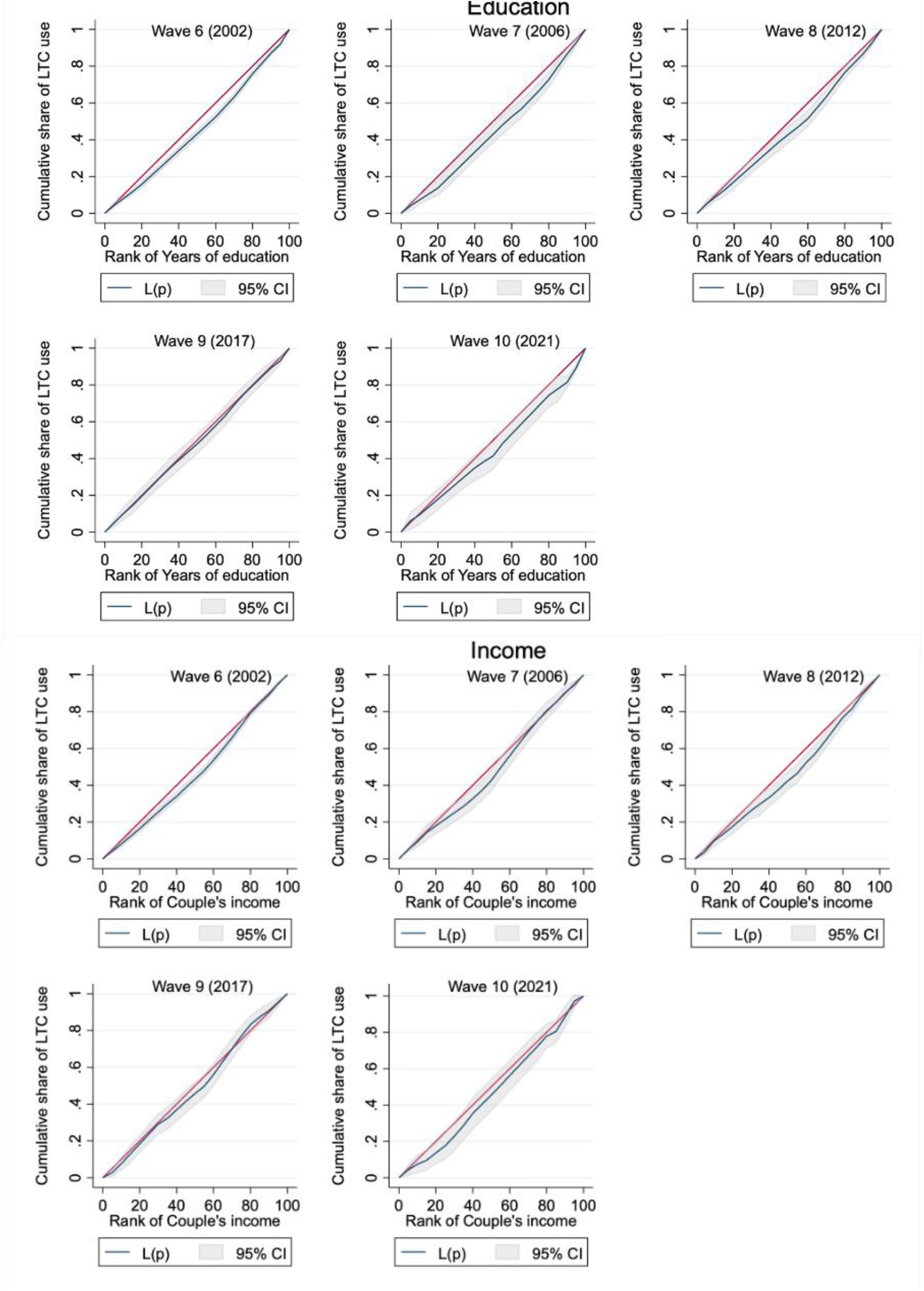
Concentration curves on standardised long-term care use by education and income. Note: The 95% confidence interval (CI; grey area) is calculated based on standard errors (SE) adjusted for clusters for each respondent; estimates are weighted by both cross-sectional and longitudinal weights. L(p) is the concentration curve for need-standardised long-term care (LTC) use.

**Table 4.** Concentration indices for medical and long-term care use.

|  |  |  | 2002 | 2006 | 2012 | 2017 | 2021 |
| --- | --- | --- | --- | --- | --- | --- | --- |
| Forgone medical care | Education | Concentration index |  |  | -0.02 | 0.11 | -0.02 |
|  |  | SE |  |  | 0.07 | 0.07 | 0.08 |
|  |  | P-value |  |  | 0.73 | 0.11 | 0.80 |
|  |  | N |  |  | 2,044 | 1,264 | 1,542 |
|  | Income | Concentration index |  |  | -0.09 | -0.00 | 0.04 |
|  |  | SE |  |  | 0.08 | 0.08 | 0.09 |
|  |  | P-value |  |  | 0.26 | 0.99 | 0.63 |
|  |  | N |  |  | 1,744 | 1,103 | 1,224 |
| Unmet need for long-term care | Education | Concentration index | -0.13 | 0.03 | -0.00 | -0.04 | 0.05 |
|  |  | SE | 0.06 | 0.06 | 0.06 | 0.07 | 0.09 |
|  |  | P-value | <0.05 | 0.60 | 0.99 | 0.55 | 0.58 |
|  |  | N | 442 | 385 | 471 | 320 | 157 |
|  | Income | Concentration index | -0.14 | -0.16 | -0.07 | -0.03 | 0.07 |
|  |  | SE | 0.06 | 0.07 | 0.07 | 0.08 | 0.12 |
|  |  | P-value | <0.05 | <0.05 | 0.30 | 0.71 | 0.54 |
|  |  | N | 376 | 300 | 381 | 256 | 118 |
| Non-standardised long-term care use | Education | Concentration index | 0.11 | 0.22 | 0.12 | -0.00 | 0.16 |
|  |  | SE | 0.06 | 0.06 | 0.06 | 0.07 | 0.10 |
|  |  | P-value | 0.08 | <0.01 | <0.05 | 0.96 | 0.09 |
|  |  | N | 442 | 385 | 471 | 320 | 157 |
|  | Income | Concentration index | 0.07 | 0.08 | 0.14 | 0.07 | 0.08 |
|  |  | SE | 0.07 | 0.07 | 0.07 | 0.08 | 0.11 |
|  |  | P-value | 0.34 | 0.25 | <0.05 | 0.42 | 0.49 |
|  |  | N | 376 | 300 | 381 | 256 | 118 |
| Standardised long-term care use | Education | Concentration index | 0.07 | 0.12 | 0.09 | -0.00 | 0.08 |
|  |  | SE | 0.04 | 0.03 | 0.03 | 0.04 | 0.05 |
|  |  | P-value | 0.05 | <0.01 | <0.01 | 0.94 | 0.13 |
|  |  | N | 442 | 385 | 471 | 320 | 157 |
|  | Income | Concentration index | 0.04 | 0.05 | 0.08 | 0.03 | 0.06 |
|  |  | SE | 0.04 | 0.04 | 0.04 | 0.04 | 0.06 |
|  |  | P-value | 0.32 | 0.17 | <0.05 | 0.43 | 0.33 |
|  |  | N | 376 | 300 | 381 | 256 | 118 |

Note: Standard errors (SE) are adjusted for clusters in each respondent; Wagstaff indices are presented; and estimates are weighted by both cross-sectional and longitudinal weights.

### Heterogeneity by sex and age

Standardised LTC use was pro-higher-education and pro-rich for both men and women (Appendix Table A-3). CI-based inequities in each wave were particularly evident among women, corresponding to the results of our primary analysis (Appendix Table A-4). Regarding the heterogeneity by age, we observed similar pro-higher education, particularly among those aged 80 years (Appendix A-5 and A-6).

## Discussion

This study aimed to assess unmet need and socioeconomic inequity in LTC access and found three main findings. First, more than one-third of the participants underwent unmet needs for LTC. Second, LTC access was pro-higher-education, which was more evident than income inequity. Third, pro- higher-educational inequity was evident among women and the oldest-old population, despite being descriptively similar among their male and younger (i.e. 60–80 years) counterparts.

Japan has the highest UHC achievement (39); nonetheless, challenges exist with certain proportions of people experiencing unmet medical and LTC needs. People with higher education may be better at accessing the necessary services, considering health literacy is related to education (40) and predicts a lack of understanding and use of health services (41). For medical care, older people might not experience unmet needs frequently due to the absence of gatekeeping and low copayment rates.

Educational inequity in LTC use was more pronounced among women and the old. This may be explained by different help-seeking behaviours by men and women despite similar health needs. Women are more likely to seek and use care than men (42); therefore, disparities could be more apparent when a specific group forgoes it.

Our findings generate policy implications for enhancing access to care. First, barriers to accessing LTC or relevant services may exist because people need to make an application by themselves. In Japan’s policies for care for older people, community comprehensive support centres located in each municipality play an important role in care management and facilitating the use of necessary care. Nevertheless, reaching out to these centres still requires care seeking from people with some difficulties themselves or their family members For some older people, a decline in intrinsic capacity and care needs may be detected by health check-ups or their attending physicians. However, poor health is associated with social isolation (43); therefore, people with health issues may be unable to access the necessary care because of physical barriers (e.g. the lack of transportation) or health-literacy-related issues (e.g. the lack of knowledge about how to seek care). Therefore, it is necessary to enhance access through any of these channels, such as education, to improve an understanding of available resources, strengthening the functions of health centres and communities to detect the needs of citizens, and facilitating their access to necessary care.

This study had four limitations. First, the sample size was small because we focused only on individuals with LTC needs. Therefore, we may not obtain significant results for some analyses or may be unable to generalise our findings. Second, our measure of LTC certification was self-reported and did not include information on the number of services used. Further studies should address these limitations by using extensive administrative data. Third, we could not distinguish between unmet needs for formal and informal LTC because of data restrictions. Depending on the policy context in each country, the availability and service coverage of formal long-term care services largely vary. Therefore, assessing the unmet needs for both formal and informal care comprehensively is required to design the service coverage of formal care. Fourth, there were non-responders, particularly in Wave 10. Thus, we may have missed vulnerable populations during the state of emergency in large cities, by which we cannot conclude that inequity was absent during the pandemic.

## Conclusions

In conclusion, approximately 42% of individuals with LTC needs experienced an unmet need for ADL or IADL support, and approximately 62% were not certified for public LTC use even with ADL or IADL limitations. Inequity in care access was driven by education rather than income. Even if the services are universally available, financial and non-financial supports are indispensable to ensuring all people can access care whenever and wherever they need them.

## Data Availability

Data until Wave 8 are available with permission to use through The Social Science Japan Data Archive.

https://ssjda.iss.u-tokyo.ac.jp/Direct/datasearch.php

## Declarations

### Ethics approval and consent to participate

This study was approved by the institutional review board of the Tokyo Metropolitan Geriatric Hospital and Institute of Gerontology (R21-008). All participants provided their informed consent to participate in the study.

### Consent for publication

Consent for publication was not required because all information collected through the surveys was deidentified.

### Availability of data and materials

The data used for this study are partially accessible through the webpage of the Centre for Social Research and Data Archives, Institute of Social Sciences of The University of Tokyo: https://csrda.iss.u-tokyo.ac.jp/english/infrastructure/

### Authors’ contributions

SO conceptualised the study, conducted statistical analysis, and prepared a first draft, which was refined by AY, EK, and JL. SO, AY, EK, and JL contributed to the design of the survey and data collection. All authors have reviewed and agreed to the final draft of the submitted paper.

## Acknowledgements

We are grateful for the helpful comments from Drs Megumi Rosenberg and Shinichi Tomioka at the World Health Organization Centre for Health Development.

## Appendix A

**Appendix Table A-1.** A list of need and non-need variables.

| Classification | Variable | Description |
| --- | --- | --- |
| Need | Age | Respondent's age |
|  | Gender | Respondent's gender |
|  | Self-rated health | 1: Very bad, 2: Bad, 3: Fair, 4: Good, 5: Very good |
|  | Chronic conditions | The number of chronic conditions respondents suffer from, including chronic respiratory diseases, hypertension, heart diseases, diabetes, stroke, and cancer (0–6) |
|  | Activities of daily living (ADL) | A sum of the magnitudes of difficulties in performing bathing, dressing, feeding, transferring, outing, and toileting, ranging from 1: Never difficult to 5: Unable to do at all (0–24) |
| Need | Instrumental activities of daily living (IADL) | A sum of the magnitudes of difficulties in shopping for personal items, using a telephone, riding the bus or subway alone, and performing light tasks around the house, ranging from 1: Never difficult to 5: Unable to do at all (0–16) |
|  | Cognitive functioning | The number of misreporting of cognitive functioning assessments comprising respondent's home address, interview date, interview day, mother's maiden name, name of the current prime minister, name of the previous prime minister, a simple calculation, respondent's birthday, and respondent's age (0–9) |
| Non-need | Marital status | 1 if single (including unmarried, divorced, and widowed); 0 otherwise |
|  | Co-residence | The number of family members living together with a respondent |
|  | Residential area (Geographic region of Japan) | Hokkaido, Tohoku, Kanto, Hokuriku, Tozan, Tokai, Kinki, Chugoku, Shikoku, and Kyushu |
|  | Municipal/population category of residential area | Government-designated, 200K+, 100K-200K, <100K, and Towns and villages |
|  | Year | Wave 6 (2002), Wave 7 (2006), Wave 8 (2012), Wave 9 (2017), Wave 10 (2021) |

**Appendix Table A-2.**
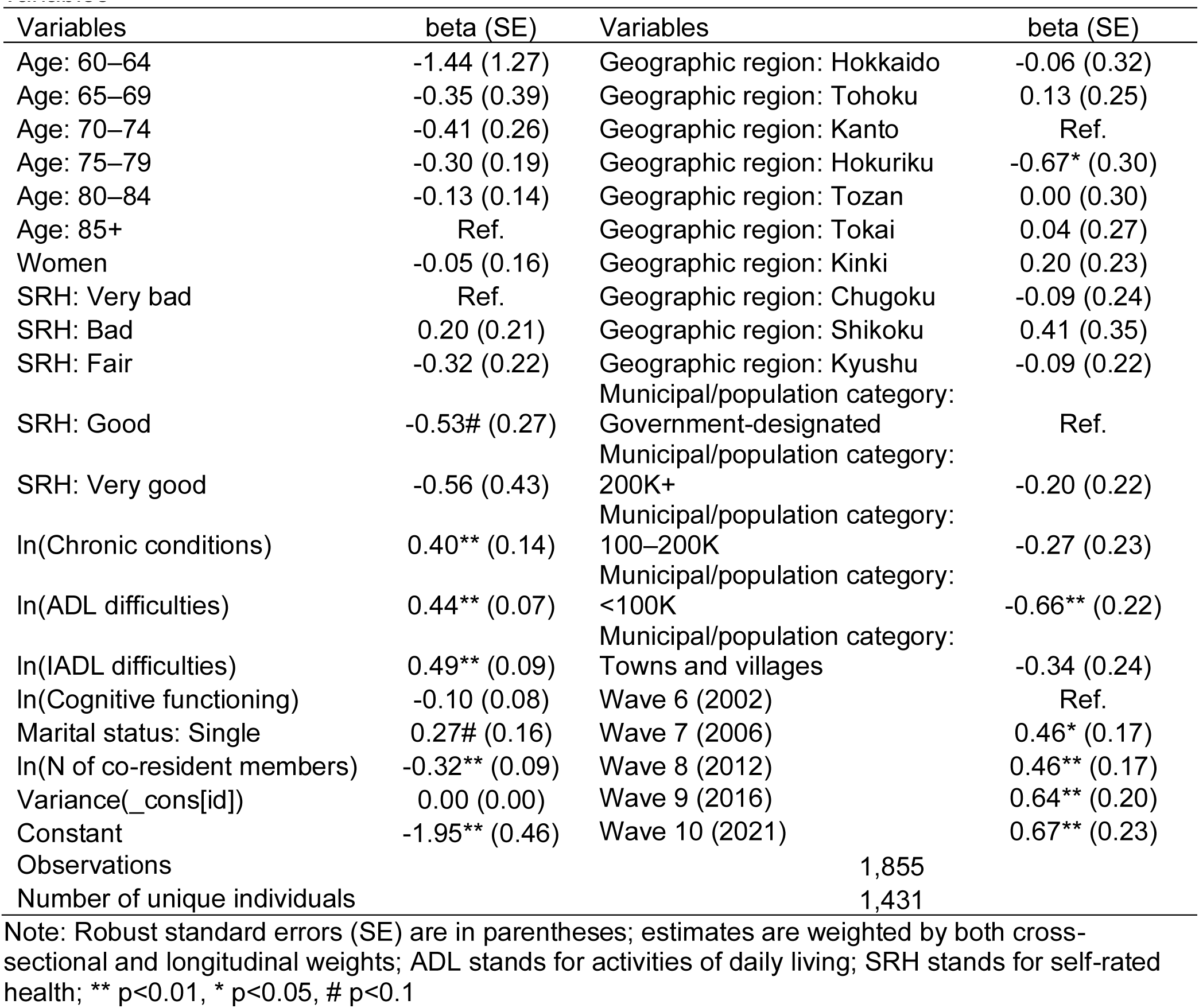
Regression results: Prediction of long-term care use by need and non-need variables.

**Table A-3.**
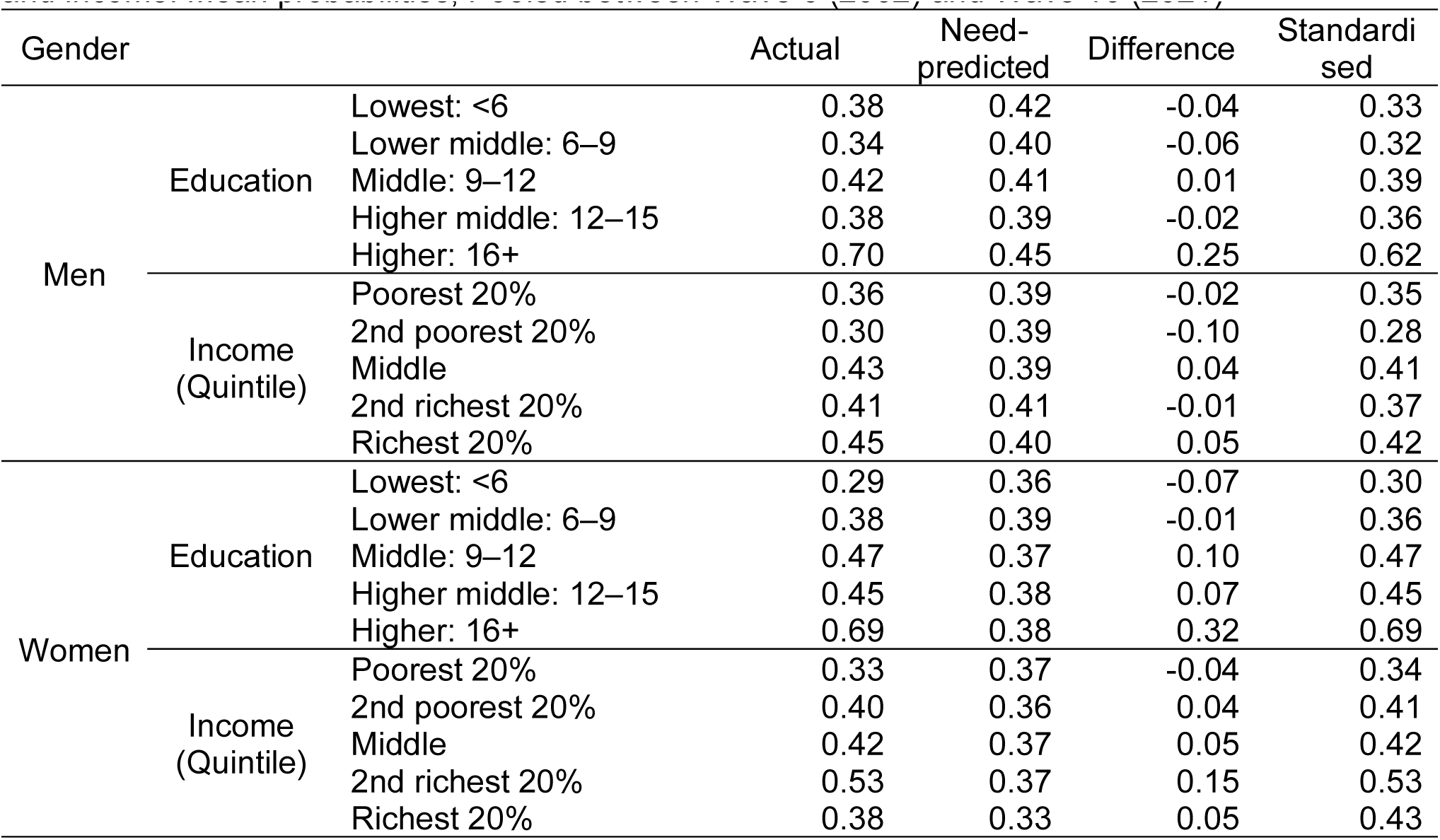
Gender-specific non-standardised and standardised long-term care use by education and income: Mean probabilities, Pooled between Wave 6 (2002) and Wave 10 (2021)

**Table A-4.** Concentration indices for long-term care use by gender.

| Gender |  |  | 2002 | 2006 | 2012 | 2017 | 2021 |
| --- | --- | --- | --- | --- | --- | --- | --- |
| Men | Education | Concentration index | 0.02 | 0.10 | 0.06 | -0.02 | 0.15 |
|  |  | SE | 0.10 | 0.06 | 0.06 | 0.07 | 0.08 |
|  |  | P-value | 0.85 | 0.10 | 0.36 | 0.79 | 0.05 |
|  |  | N | 130 | 103 | 135 | 114 | 57 |
|  | Income | Concentration index | -0.11 | 0.12 | 0.10 | 0.00 | -0.06 |
|  |  | SE | 0.09 | 0.07 | 0.06 | 0.07 | 0.08 |
|  |  | P-value | 0.21 | 0.09 | 0.12 | 0.94 | 0.49 |
|  |  | N | 120 | 79 | 119 | 99 | 46 |
| Women | Education | Concentration index | 0.10 | 0.14 | 0.11 | 0.02 | 0.02 |
|  |  | SE | 0.04 | 0.04 | 0.04 | 0.05 | 0.07 |
|  |  | P-value | <0.01 | <0.01 | <0.01 | 0.64 | 0.76 |
|  |  | N | 312 | 282 | 336 | 206 | 100 |
|  | Income | Concentration index | 0.11 | 0.03 | 0.08 | 0.06 | 0.11 |
|  |  | SE | 0.04 | 0.04 | 0.05 | 0.05 | 0.08 |
|  |  | P-value | <0.01 | 0.42 | 0.07 | 0.31 | 0.18 |
|  |  | N | 256 | 221 | 262 | 157 | 72 |

Note: Standard errors (SE) are adjusted for clusters in each respondent; Wagstaff indices are presented; and estimates are weighted by both cross-sectional and longitudinal weights.

**Table A-5.** Age-specific non-standardised and standardised long-term care use by education and income: Mean probabilities, Pooled between Wave 6 (2002) and Wave 10 (2021)

| Age |  |  | Actual | Need-predicted | Difference | Standardised |
| --- | --- | --- | --- | --- | --- | --- |
| 60–79 | Education | Lowest: <6 | 0.27 | 0.31 | -0.04 | 0.33 |
|  |  | Lower middle: 6–9 | 0.32 | 0.37 | -0.05 | 0.32 |
|  |  | Middle: 9–12 | 0.36 | 0.34 | 0.02 | 0.40 |
|  |  | Higher middle: 12–15 | 0.32 | 0.40 | -0.07 | 0.30 |
|  |  | Higher: 16+ | 0.59 | 0.39 | 0.20 | 0.57 |
|  | Income (Quintile) | Poorest 20% | 0.33 | 0.35 | -0.02 | 0.36 |
|  |  | 2nd poorest 20% | 0.37 | 0.38 | -0.01 | 0.36 |
|  |  | Middle | 0.46 | 0.33 | 0.12 | 0.50 |
|  |  | 2nd richest 20% | 0.30 | 0.34 | -0.04 | 0.33 |
|  |  | Richest 20% | 0.32 | 0.32 | 0.00 | 0.37 |
| 80+ | Education | Lowest: <6 | 0.31 | 0.39 | -0.07 | 0.30 |
|  |  | Lower middle: 6–9 | 0.39 | 0.40 | -0.02 | 0.36 |
|  |  | Middle: 9–12 | 0.50 | 0.40 | 0.10 | 0.47 |
|  |  | Higher middle: 12–15 | 0.47 | 0.38 | 0.09 | 0.46 |
|  |  | Higher: 16+ | 0.78 | 0.48 | 0.30 | 0.67 |
|  | Income (Quintile) | Poorest 20% | 0.34 | 0.38 | -0.04 | 0.33 |
|  |  | 2nd poorest 20% | 0.13 | 0.29 | -0.16 | 0.21 |
|  |  | Middle | 0.41 | 0.39 | 0.02 | 0.40 |
|  |  | 2nd richest 20% | 0.53 | 0.40 | 0.13 | 0.51 |
|  |  | Richest 20% | 0.47 | 0.39 | 0.08 | 0.45 |

**Appendix Table A-6.** . Concentration indices for long-term care use by age.

| Age |  |  | 2002 | 2006 | 2012 | 2017 | 2021 |
| --- | --- | --- | --- | --- | --- | --- | --- |
| 60–79 | Education | Concentration index | 0.01 | 0.15 | 0.02 | -0.13 | 0.08 |
|  |  | SE | 0.05 | 0.06 | 0.07 | 0.18 | 0.07 |
|  |  | P-value | 0.76 | <0.05 | 0.75 | 0.48 | 0.29 |
|  |  | N | 207 | 118 | 102 | 33 | 58 |
|  | Income | Concentration index | 0.02 | 0.00 | 0.02 | 0.02 | 0.04 |
|  |  | SE | 0.05 | 0.06 | 0.08 | 0.16 | 0.09 |
|  |  | P-value | 0.65 | 0.98 | 0.78 | 0.89 | 0.65 |
|  |  | N | 184 | 98 | 80 | 28 | 43 |
| 80+ | Education | Concentration index | 0.13 | 0.12 | 0.13 | 0.03 | 0.06 |
|  |  | SE | 0.05 | 0.04 | 0.04 | 0.04 | 0.08 |
|  |  | P-value | <0.05 | <0.01 | <0.01 | 0.37 | 0.43 |
|  |  | N | 235 | 267 | 369 | 287 | 99 |
|  | Income | Concentration index | 0.07 | 0.09 | 0.11 | 0.04 | 0.09 |
|  |  | SE | 0.07 | 0.05 | 0.04 | 0.04 | 0.07 |
|  |  | P-value | 0.35 | 0.07 | <0.01 | 0.37 | 0.19 |
|  |  | N | 192 | 202 | 301 | 228 | 75 |

Note: Standard errors (SE) are adjusted for clusters in each respondent; Wagstaff indices are presented; and estimates are weighted by both cross-sectional and longitudinal weights.

**Figure A-1.**
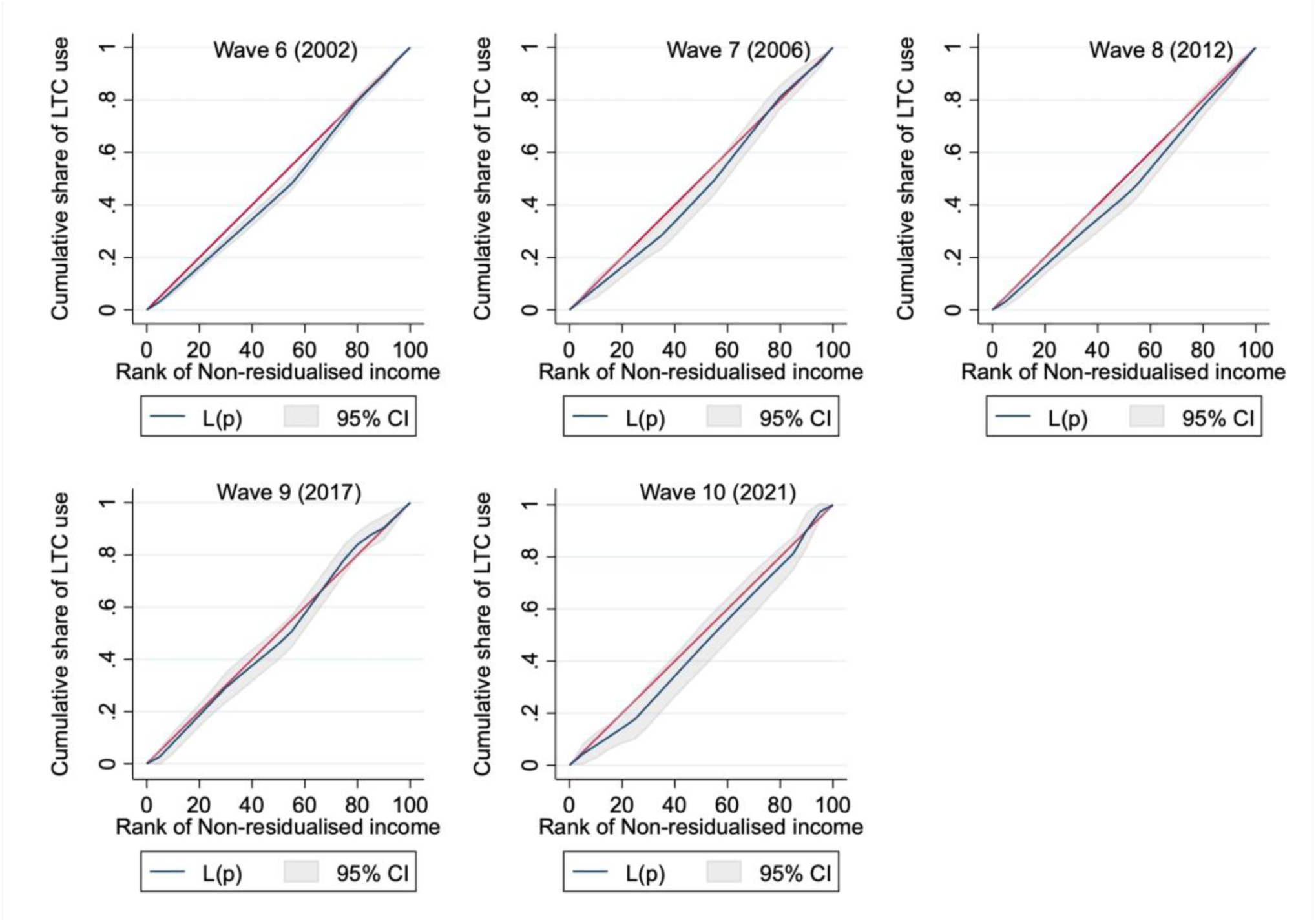
Concentration curves on standardised long-term care use by non-residualised income. Note: The 95% confidence interval (CI) is calculated based on standard errors (SE) adjusted for clusters for each respondent; estimates are weighted by both cross-sectional and longitudinal weights. The sample sizes in each wave were 376, 300, 382, 256, and 118, respectively, and L(p) is the concentration curve for long-term care (LTC).

## Appendix B: Income Residualisation

Employed and unemployed individuals are non-comparable because of large differences: theoretically, people decumulate their savings to smooth their consumption levels, particularly after retirement (i.e. life-cycle hypothesis) (1); Thus, a low income does not always suggest being deprived among older people. Instead of using limited information about savings and consumption from the National Survey of the Japanese Elderly, we residualised the income to partially adjust for the differences between workers and non-workers by fixed-effects ordinary least squares as follows:

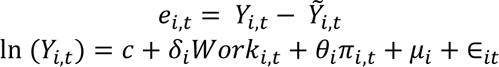

where *e*_*i*,*t*_ is the residualised income of individual i in year t, defined as the predicted income (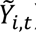) subtracted from the actual income (*Y*_*i*,*t*_). The income is predicted by the employment status (*Work*_*i*,*t*_; =1 if in paid work; =0 if otherwise) and other independent variables (*π*_*i*,*t*_), including the age, sex, marital status, residential area, municipal or population category of residential area, year-fixed effects, and individual fixed effects (*μ*_*i*_). *c* is a constant, *ε* is the stochastic disturbance, and *δ* and *θ* are the estimated coefficients of the independent variables. This formalisation enabled controlling for potential imbalances in the income arising from the employment status, demographic factors, price differences across areas and years, and time-invariant individual heterogeneity (Appendix Table B-1).

**Appendix Table B-1.** Regression result: Prediction of couple’s income.

| Variables | beta (SE) |
| --- | --- |
| Age: 60–64 | Ref. |
| Age: 65–69 | -0.02 (0.02) |
| Age: 70–74 | -0.01 (0.03) |
| Age: 75–79 | -0.04 (0.05) |
| Age: 80–84 | -0.04 (0.05) |
| Age: 85+ | -0.05 (0.08) |
| Employment status: Currently working | 0.20** (0.02) |
| Marital status: Single | -0.06* (0.02) |
| Individual FE | Yes |
| Residential area FE | Yes |
| City-scale FE | Yes |
| Year FE | Yes |
| Constant | 5.13**<br>(0.03) |
| Observations | 17,031 |
| Number of unique individuals | 4,586 |

Note: Robust standard errors (SE) are in parentheses; estimates are weighted by both cross-sectional and longitudinal weights; FE denotes fixed effects; \*\* $p < 0.01$ , \* $p < 0.05$ , # $p < 0.1$

